# Survey of nationwide public perceptions regarding acceptance of wastewater used for community health monitoring in the United States

**DOI:** 10.1101/2022.03.16.22272262

**Authors:** A. Scott LaJoie, Rochelle H. Holm, Lauren B. Anderson, Heather D. Ness, Ted Smith

**Affiliations:** Department of Health Promotion and Behavioral Sciences, School of Public Health and Information Sciences, University of Louisville, 485 E. Gray St., Louisville, KY 40202, United States; Christina Lee Brown Envirome Institute, School of Medicine, University of Louisville, 302 E. Muhammad Ali Blvd., Louisville, KY 40202, United States

**Keywords:** community health, environmental monitoring, Privacy Attitudes Questionnaire, public opinion, sewer, wastewater-based epidemiology

## Abstract

During the coronavirus disease 2019 (COVID-19) pandemic researchers looked for evidence of severe acute respiratory syndrome coronavirus 2 (SARS-CoV-2) RNA in feces dissolved in wastewater samples to assess levels of infection across communities. This activity is known colloquially as sewer monitoring and called wastewater-based epidemiology in academic settings. When used for public health surveillance in the United States, wastewater monitoring is not regulated, although general ethical principles have been described. Prior to this study, no nationwide data existed regarding the public’s perceptions of wastewater being used for community health monitoring. Using an online survey distributed to a representative sample of adults in the Unites States (N=3,083), we investigated the public’s perceptions regarding what is monitored, where monitoring occurs, and privacy concerns related to wastewater monitoring as a public health surveillance tool. Further, the Privacy Attitudes Questionnaire assessed respondents’ general privacy boundaries. The results suggest that respondents supported using wastewater for health monitoring, but within some bounds. Participants were most likely to support or strongly support monitoring for disease (95%), environmental toxins (94%), and terroristic threats (90%, e.g., anthrax). Two-thirds of respondents endorsed no prohibition to locations being monitored while the most common category of location respondents wanted to be prohibited from monitoring was personal residencies. Additionally, the findings suggest that those younger in age and living in an urban area were more supportive of wastewater monitoring, compared to older, suburban dwellers.

## 1. Introduction

Wastewater monitoring for public health surveillance is a tool that detects biological and chemical targets in sewage from residential or institutional settings prior to treatment and release into the environment. Typically, this involves collecting samples from the existing piped wastewater infrastructure (sewers) and has been deployed for a range of public health inquiries including tracking of pathogens [1], illicit drugs [2,3], dietary patterns [4], and biological agents as weapons [5]. During the coronavirus disease 2019 (COVID-19) pandemic researchers looked for evidence of severe acute respiratory syndrome coronavirus 2 (SARS-CoV-2) RNA in feces dissolved in wastewater samples to assess levels of infection across communities [6-8]. Sewer monitoring to assess the incidence, distribution, and possible control of diseases is often called wastewater-based epidemiology (WBE). During the COVID-19 pandemic, much discussion has occurred about sewer monitoring potentially serving as the foundation for innovative and cost-effective methods for public health action and policy broadly. However, surveillance activities can evoke privacy concerns and possible stigmatization of institutional settings or communities where concerning levels of health risks are identified. Current WBE methods do not involve the study of human DNA markers that may be present in the sewage; therefore, the public debate around discarded DNA does not currently apply here [9].

In the United States, WBE is not regulated regarding privacy concerns, though globally general ethical principles have been described based on the premise that samples are typically collected with permission from a utility operating through publicly owned infrastructure [10-14]. Pertinently, the premise of WBE is that informed and voluntary consent to participate in wastewater monitoring is not needed from individuals contributing feces or urine to the wastewater sample [10-14]. Most wastewater utilities in the United States are governed by public utility commissions which are charged with serving the public interest. Yet, to date, there have been no national assessments of WBE when used for public health surveillance to determine public acceptance or concern.

Technologies that use impersonal data for a service purpose such as civil status (birth, death, and marriage), housing, elections, or work, have been shown to less likely raise privacy concerns [15]. In contrast, technologies that use personal data for surveillance purposes such as police data or images captured by closed-circuit television cameras are more likely to raise privacy concerns [15]. In this regard, there are three recurring dimensions: sensitivity/personalness of the data, purpose (service versus surveillance) of the data collection, and the collector/user of the data [15]. Each of these three dimensions can be extended to the application of wastewater monitoring. For instance, legislation opposing COVID-19 WBE include North Dakota’s House Bill 1348, which was aimed at “prohibiting the testing of wastewater for genetic material or evidence of disease; and to provide a penalty,” did not pass in February 2021 [16]. Media reporting around this proposed legislation focused on the surveillance purposes per privacy concerns of building-level surveillance, stating that the practice could violate college student’s privacy rights [17]. During the COVID-19 pandemic, the shifting policies of social restrictions determined by community infection levels created circumstances that made credible concerns about WBE data use as partial evidence for changing societal conduct such as community, school, or industry lockdown conditions.

Although WBE has been well established to assess public health [1-5], the COVID-19 pandemic magnified the field and may have increased public awareness accordingly. Using a survey distributed to a representative sample of adults in the Unites States, we investigated the awareness, acceptance, and privacy concerns related to wastewater monitoring as a public health surveillance tool. The aim of this study was to assess: (1) acceptance and awareness of wastewater monitoring; (2) knowledge and perceptions of privacy issues; and (3) factors that influence an individual’s level of awareness and acceptance of wastewater monitoring. The results will be used to develop insights regarding the acceptability of monitoring and inform policies regarding future applications of wastewater monitoring at both national and local levels.

## 2. Methods and Materials

An online survey was distributed to a representative sample of adults in the Unites States in January and February 2022. Survey respondents were 18 years and older, residents of the United States, and registered with the Qualtrics XM (Provo, UT) participant panel. Additional inclusion criteria included the ability to read and understand the English language and self-reported to live in an area considered urban or suburban. Potential participants were contacted by email invitation. Invitations were sent randomly to a pool of participants who met the study’s inclusion criteria and who had signed up previously to receive study invitations from Qualtrics.

Study participants were directed to a secure website reviewed by an institutional review board that complied with the Health Insurance Portability and Accountability Act of 1996 (HIPAA), where they were shown a preamble informed consent. Completion of the survey, which included no identifiable or protected information, was considered assent to participate.

### 2.1. Data collection instrument

The 80-item survey included three components: (1) questions to assess knowledge, awareness, and acceptance of wastewater monitoring; (2) questions covering demographics (gender identity, race, ethnicity, age, income, education level, geography); and (3) questions on privacy concerns using the Privacy Attitudes Questionnaire (PAQ) [18]. The PAQ was used as way to structure privacy as a psychometric construct and considers privacy being limited in both the physical and digital public environment.

Within the survey, several items were written to theoretically cluster into subscales. Reliability analysis provides evidence the survey was interpreted and used as intended. Subscales included: *knowledge of public health activities* (n=6, α=0.86), *support for sewage monitoring of activities* (n=10, α=0.87), *support for monitoring of locations* (n=7, α=0.84), *opposition of monitoring of types of locations* (n=6, α=0.84), and the *Privacy Attitude Questionnaire* (n=37, α=0.60). According to its developers, the PAQ produces results in four domains. In this study, the reliability estimates of these four domains were: *exposure* (n=9, α=0.54), *monitored* (n=9, α=0.39), *personal information* (n=10, α=0.70), and *protection* (n=8, α=0.69).

Additional items assessed *knowledge* (n=3), *self vs. other orientation* (n=6), and *confidence and willingness to share personal information* (n=6). Responses to three subscales showed reliability estimates that were less acceptable (αs less than 0.5). Demographics (n=8) were collected to estimate whether the results could be generalized to the United States population.

Survey items consisted of Likert-type scales (arranged in matrices), rank-ordering, select-one, and choose-all that apply.

### 2.2. Data management and analysis

Online surveys using participant panels pose a risk of false responding (e.g., providing random errors to earn an incentive). To counter this risk, the survey was administered via the company Qualtrics XM to participants who are regularly screened for fidelity. Qualtrics XM distributes the survey invitations randomly to its pool of participants who meet inclusion criteria, performs an initial scrubbing of the data, and compensates the participants whose data is deemed acceptable by standards set by Qualtrics XM. These standards include satisfying a CAPTCHA test to access the survey, time spent to complete the survey, missing data analysis, and additional proprietary algorithms. Data not meeting Qualtrics XM standards is excluded. The present data collection resulted in 386 (11%) of responses being rejected. Two items were embedded in the survey as attention checks; 100% of respondents included in the analysis answered both attention checks correctly.

Given the nature of categorical data, analyses were mostly restricted to descriptive measures and non-parametric tests, including frequency counts, cross-tabulations, chi-square, or Fisher exact tests. Pseudo-continuous variables were created where appropriate (i.e., subscales using the same measure type and having a Chronbach Alpha greater than 0.6). The resulting sample size included 3,083 respondents from across the United States (Figure 1) with an estimated margin of error of +/- 2%.

**Fig 1.**
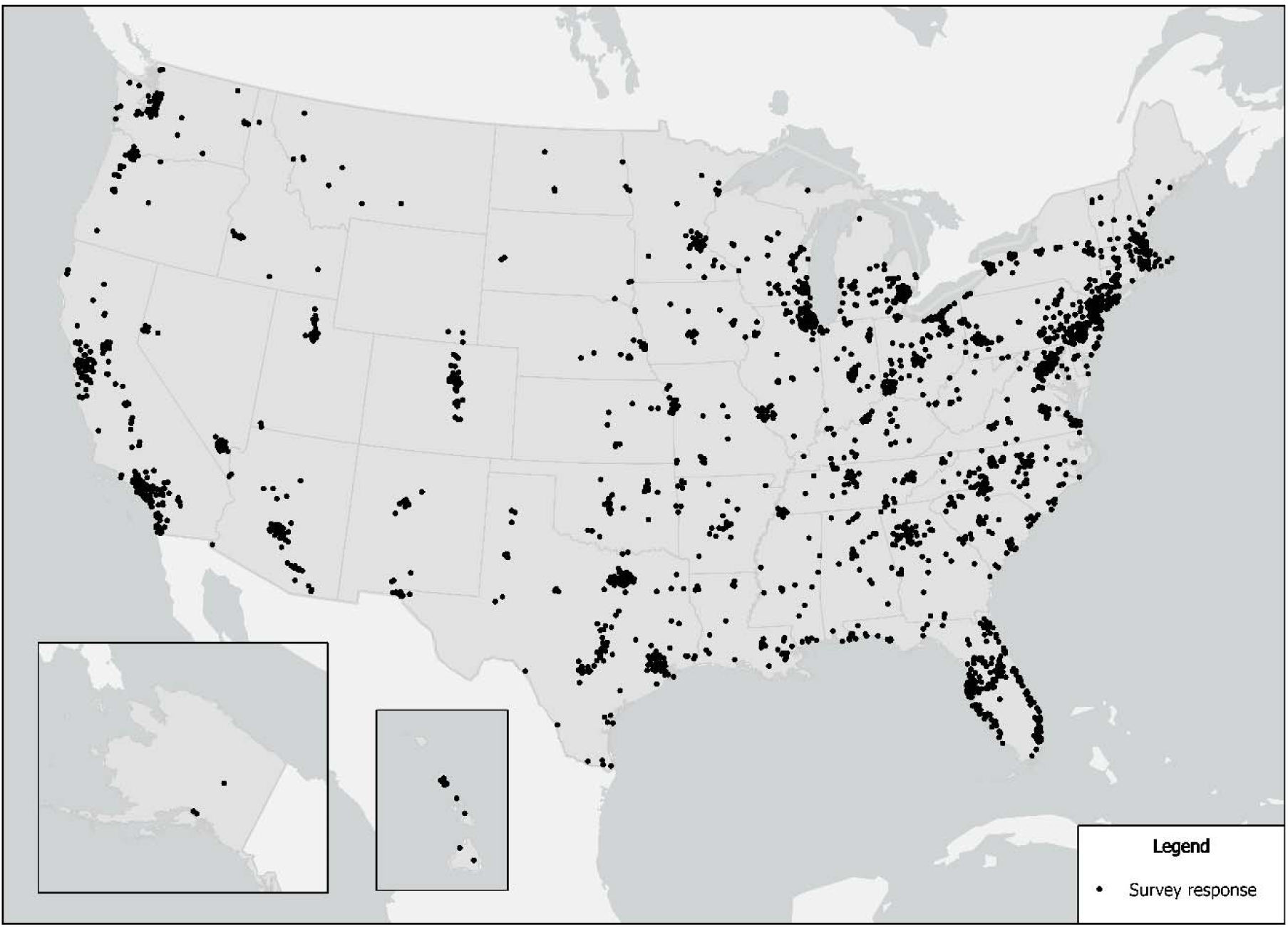
Location of respondents.

The researchers considered an alpha of <0.05 statistically significant. The data analysis for this study was generated using IBM Statistical Package for the Social Sciences (SPSS) software (version 28; Chicago, IL) [19].

### 2.3. Ethics

The University of Louisville Institutional Review Board approved this project as Human Subjects Research (IRB number: 21.0877).

## 3. Results

We obtained complete responses from 3,083 people. Respondents were mostly female (69%), white (84%), non-Hispanic (95%) and older (66% of respondents were older than 54 years). Income distribution skewed towards higher income brackets, with roughly 25% having incomes between $20,000 and $40,000; 28% between $40,000 and $70,000; and 32% having incomes greater than $70,000. The sample was largely well educated, with many (79%) having some college or beyond. Most respondents self-reported living in a suburban area (70%) compared to urban area (30%). See Table 1 for a full description of the sample.

**Table 1.**
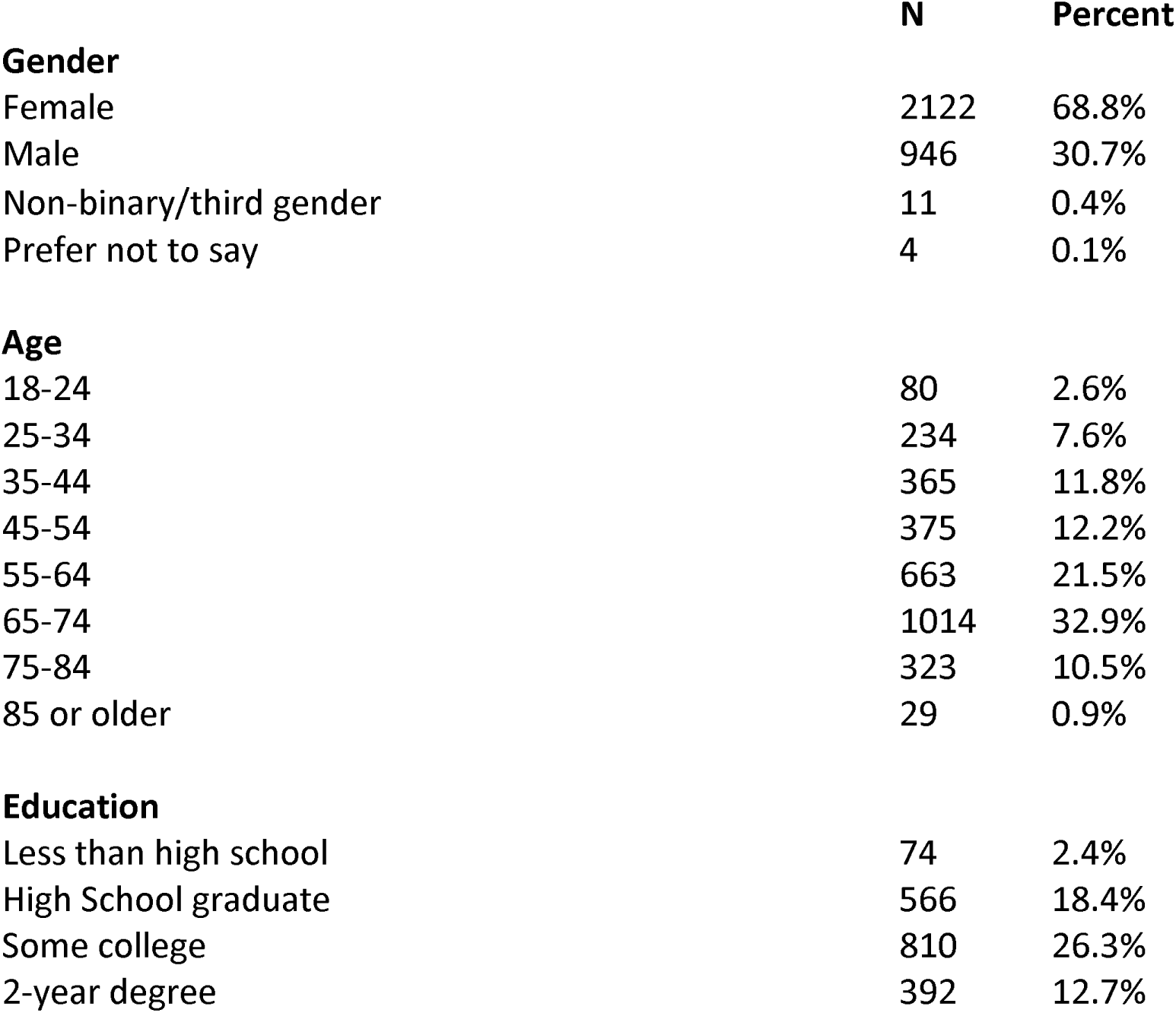
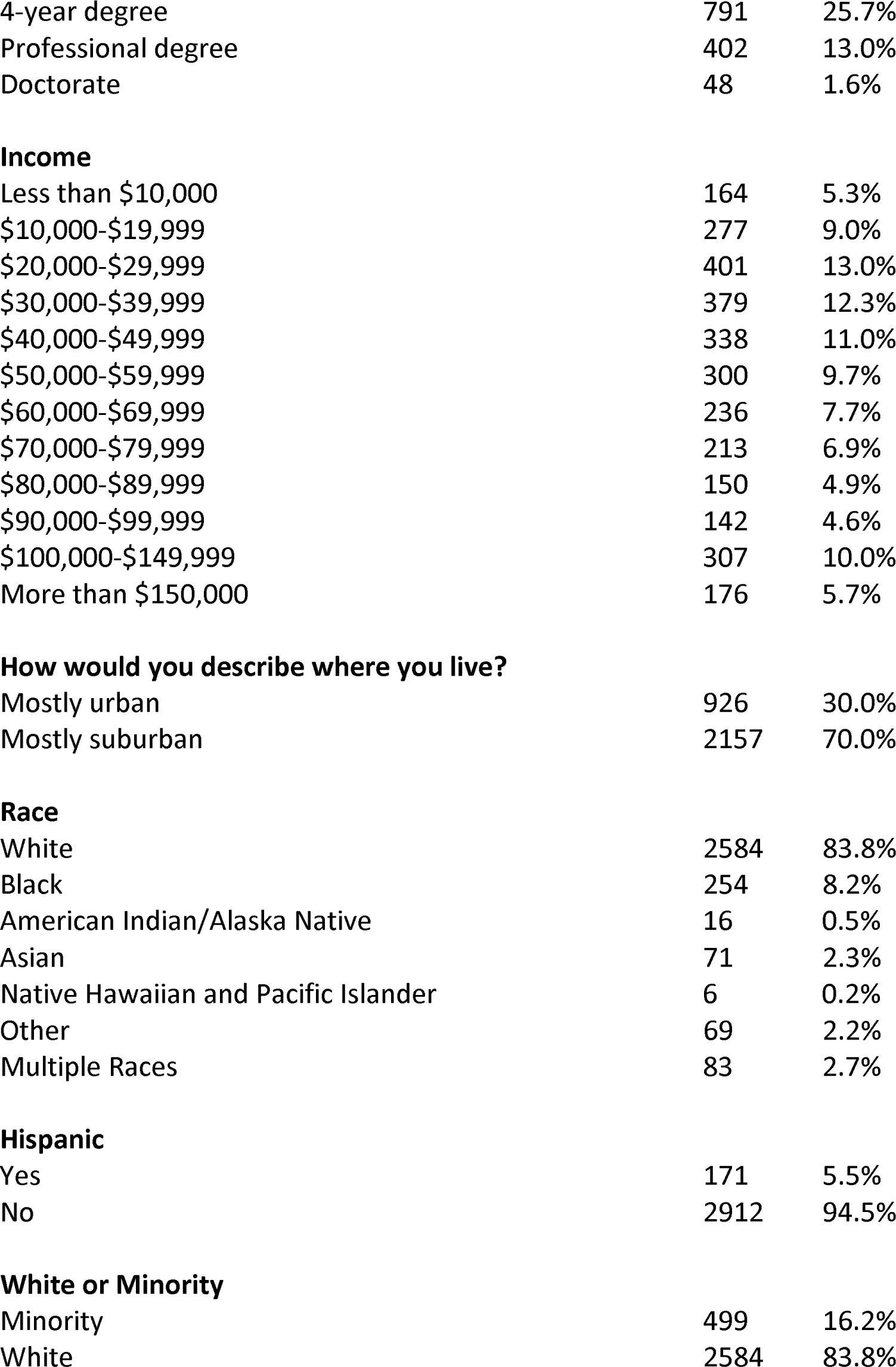
Demographic characteristics of respondents.

|  | **N** | **Percent** |
| --- | --- | --- |
| **Gender** |  |  |
| Female | 2122 | 68.8% |
| Male | 946 | 30.7% |
| Non-binary/third gender | 11 | 0.4% |
| Prefer not to say | 4 | 0.1% |
| **Age** |  |  |
| 18-24 | 80 | 2.6% |
| 25-34 | 234 | 7.6% |
| 35-44 | 365 | 11.8% |
| 45-54 | 375 | 12.2% |
| 55-64 | 663 | 21.5% |
| 65-74 | 1014 | 32.9% |
| 75-84 | 323 | 10.5% |
| 85 or older | 29 | 0.9% |
| **Education** |  |  |
| Less than high school | 74 | 2.4% |
| High School graduate | 566 | 18.4% |
| Some college | 810 | 26.3% |
| 2-year degree | 392 | 12.7% |
| 4-year degree | 791 | 25.7% |
| Professional degree | 402 | 13.0% |
| Doctorate | 48 | 1.6% |
| **Income** |  |  |
| Less than \$ 10,000 | 164 | 5.3% |
| \$10,000-\$19,999 | 277 | 9.0% |
| \$20,000-\$29,999 | 401 | 13.0% |
| \$30,000-\$39,999 | 379 | 12.3% |
| \$40,000-\$49,999 | 338 | 11.0% |
| \$50,000-\$59,999 | 300 | 9.7% |
| \$60,000-\$69,999 | 236 | 7.7% |
| \$70,000-\$79,999 | 213 | 6.9% |
| \$80,000-\$89,999 | 150 | 4.9% |
| \$90,000-\$99,999 | 142 | 4.6% |
| \$100,000-\$149,999 | 307 | 10.0% |
| More than \$150,000 | 176 | 5.7% |
| **How would you describe where you live?** |  |  |
| Mostly urban | 926 | 30.0% |
| Mostly suburban | 2157 | 70.0% |
| **Race** |  |  |
| White | 2584 | 83.8% |
| Black | 254 | 8.2% |
| American Indian/Alaska Native | 16 | 0.5% |
| Asian | 71 | 2.3% |
| Native Hawaiian and Pacific Islander | 6 | 0.2% |
| Other | 69 | 2.2% |
| Multiple Races | 83 | 2.7% |
| **Hispanic** |  |  |
| Yes | 171 | 5.5% |
| No | 2912 | 94.5% |
| **White or Minority** |  |  |
| Minority | 499 | 16.2% |
| White | 2584 | 83.8% |

**Table 2.**
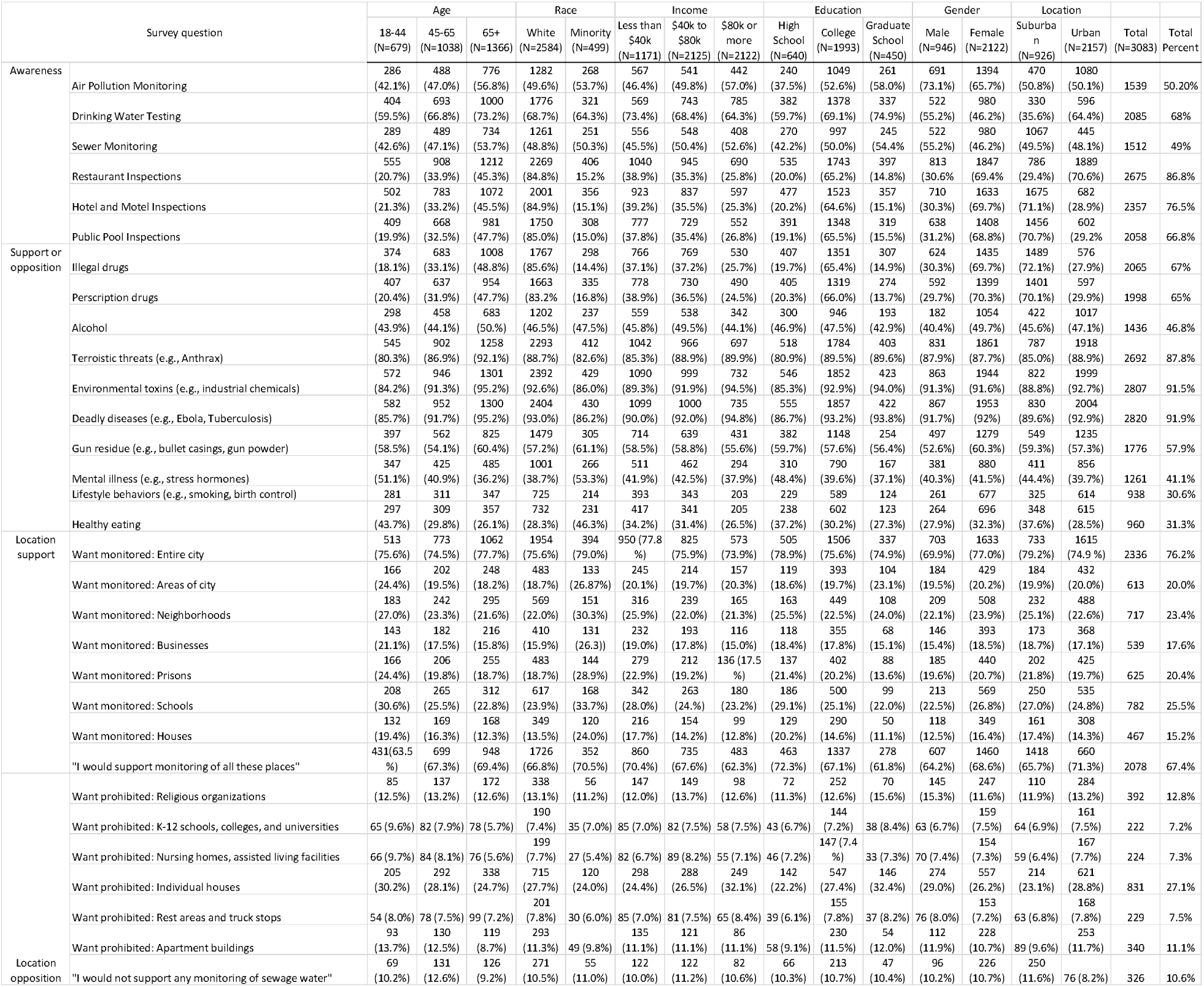
Awareness and support (N=3,083) by age, race, income, education, gender, and location.

### 3.1. Descriptive Findings

Three items assessed rudimentary knowledge of wastewater monitoring of SARS-CoV-2, the virus which causes COVID-19. Participants were asked whether COVID-19 could be detected in sewage. The correct answer (true) was selected by 1,304 (42%), while 290 (9%) chose incorrectly (false), and 1489 (48%) indicated they did not know. The next item asked which if any statement was false. There were five statements plus a “None are False” statement. The correct false answer, monitoring sewage can determine which person or persons in a household has COVID-19, was most commonly chosen (n=1,473, 48%), with 1,060 (34%) incorrectly saying none of the statements are false. Next, participants were asked to identify the fastest way to detect COVID-19 in a community. Response options included items such as test everyone, survey people, and count visits to the emergency department. The correct answer, “measure the level of the virus in the sewer water,” was chosen by 1,174 (38%). A summary knowledge score was created, with respondents earning one point for each correct answer. Possible knowledge scores ranged between 0 and 3. The distribution of respondents’ knowledge scores was: 0 (871, 28%), 1 (996, 32%), 2 (693, 23%), or 3 (523, 17%). The mean (standard deviation) was 1.28 (1.05).

Respondents were asked to rate their level of awareness of six functions of the health department on a scale of 0 (no awareness) to 4 (full awareness). Participants were mostly aware of restaurant inspections (96%), hotel and motel inspections (83%), and drinking water (74%) and pool (73%) water quality testing, but less aware that health departments monitor air quality (55%) or wastewater (53%). The mean level of awareness across the six functions was 2.8.

When asked how strongly they would support or oppose wastewater monitoring among ten indicators of human activity or health, respondents strongly opposed, opposed, or were indifferent to monitoring of lifestyle behaviors (69%; e.g., smoking, use of birth control), diets (68%), and indicators of mental illness (58%; e.g., stress hormones). Monitoring of illegal or prescription drugs, alcohol, and gun residue was supported by half to two-thirds of respondents. Finally, participants were most likely to support or strongly support monitoring for disease (95%), environmental toxins (94%), and terroristic threats (90%, e.g., anthrax). Overall, the mean level of support for these various indicators, on a 1 to 5 scale where five is strongly support, was 3.7.

In two blocks of items, respondents were asked if they would *want* or would *prohibit* monitoring specific geographic scales (ex: neighborhood or city scales) and specific types of locations. These items were presented as a check-all that apply. Nearly 90% of respondents agreed they wanted at least some areas monitored. Specifically, 76% wanted the entire city monitored. If not the entire city, respondents wanted schools (29%), neighborhoods (26%), and prisons (23%) monitored. Less support was evident for certain areas of the city (22%), businesses (20%), or houses (17%). There were specifically named locations which some respondents thought should be prohibited from monitoring: individual households (27%), houses of worship and/or religious organizations (13%), and apartment buildings (11%). Approximately 7-8% wanted to prohibit truck stops and rest areas, school campuses (K-12 and colleges), and nursing homes or assisted living facilities from monitoring. Overall, 67% of respondents would not prohibit monitoring of any of these sites.

Wastewater monitoring has the potential to gather data which some people may prefer to keep private. Respondents were asked if they had confidence ‘city officials’ could maintain the privacy of three types of personal information (health/medical, lifestyle/behaviors, financial) (no confidence to complete confidence, 0 to 4). Eighty-six percent were confident or very confident the city would keep these types of information confidential. Lifestyle and behavioral information were the area in which the highest percentage of respondents were unsure or lacked confidence (18%), followed by financial information (17%), and health/medical information (16%). When asked whether they would be willing to give up privacy (none to all, on a scale of 0 to 4) to ensure people in the community could live safe and healthy lives, 78% of respondents reported being willing to give up most or all of the three information types, with willingness to give up financial information being least frequently endorsed (45%).

The Privacy Attitude Questionnaire (PAQ) (Figure 2) included further general privacy boundaries for items such as “I would like a high fence in my backyard” and “Insurance companies should not have access to people’s health records.” The 37-item measure, with Likert scales (1-5, 5=strongly agree), clusters into four factors. Aggregate mean (standard deviation) scores of the four scales were: *exposure*=2.73 (0.55), *monitored*=3.34 (0.48), *protection*=3.92 (0.58), and *personal information*=2.36 (0.58). Our sampled population had a greater concern about sharing their personal information compared to the other three factors.

**Fig 2.**
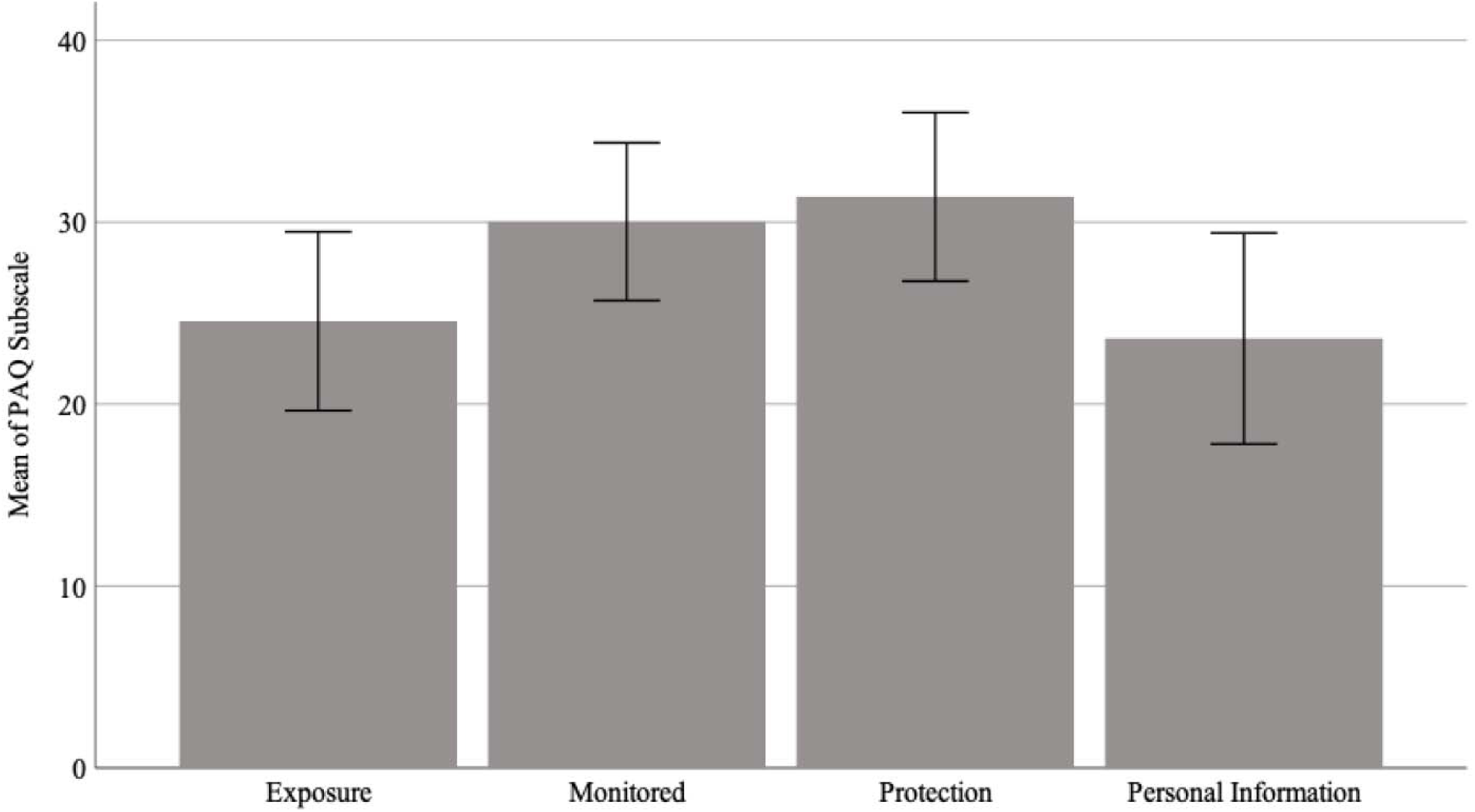
Responses from Privacy Attitudes Questionnaire. A lower score indicates a higher level of privacy concern within each factor; error bars represent standard error.

### 3.2. Inferential Findings

To understand whether differences existed in awareness, knowledge, and preferences for monitoring, several comparisons were made with the demographic variables (gender, race, age cohort, education level, income bracket, and urban/rural residency) as dependent variables. Univariate analyses of variance and t-tests were used as appropriate. Scheffe’s test was done to determine if any post-hoc comparisons were significant.

#### Knowledge

The knowledge score (mean, ranged 0 to 3) was tested for differences associated with the demographic variables. Statistical differences were found for race (*p*<0.001), age group (*p*=0.001), schooling (*p*=0.001), and income bracket (*p*=0.009); there were no differences by gender *(p*=0.71) or residency (*p*=0.38). Measures of central tendency by demographics (mean, standard deviation) were: race [white (1.33, 1.06), minority (1.02, 0.96)], age cohorts [youngest (1.10, 1.01), middle (1.23, 1.06), oldest (1.40, 1.05)], education [High School (0.95, 0.98), college (1.31, 1.05), graduate school (1.61, 1.04)], income bracket [lowest (1.14, 1.04), middle (1.33, 1.04), highest (1.44, 1.05)], gender [males (1.34, 1.04), female (1.26, 1.06)], residency [(urban (1.18, 1.05), suburban (1.33, 1.05)].

#### Awareness

Univariate analysis of variance was used to explore the main effects of the demographic variables on the average level of awareness of six public health surveillance activities. Higher means equal higher levels of awareness of the activities. The main effects of **gender** (*F*(1, 3,067)=4.13, *p*=0.047), **age cohort** (*F*(2, 3,067)=11.14, *p*<0.001), and **education** (*F*(2, 3,067)=10.97, *p*<0.001) were significant, while the main effects of residency, income bracket, or education were not (*ps*>=0.07). Specifically, males reported higher awareness (*M*=2.89, *SD*=0.91) than females (*M*=2.75, *SD*=0.95). The youngest cohort (18-44 years, *n*=679) had the lowest awareness (*M*=2.64, *SD*=0.97), followed by the middle cohort (45-64 years, *n*=1038; *M*=2.77, *SD*=0.93), with the oldest cohort (65-85+ years, *n=*1366; *M*=2.90, *SD*=0.92) being most aware. Post-hoc comparisons of age cohorts were significantly different. Those with the lowest level of education completed (high school or less, *n*=640) had the lowest awareness (*M*=2.60, *SD*=0.99) compared to those with at least some college (*n*=1993; *M*=2.83, *SD*=0.93) or graduate school (*n*=450; *M*=2.93, *SD*=0.88). Those with college and graduate school education were not different in level of awareness.

Considering the awareness of monitoring of wastewater, a similar pattern of differences was observed. Respondents who are female, older, wealthier, and more educated were more aware of wastewater monitoring. There were no differences by race or residency.

#### Support for Monitoring of Ten Activities

Respondents indicated their strength of support for the monitoring of ten activities: use of illegal and prescription drugs or alcohol, eating habits, lifestyle behaviors, gun residue, toxins, terroristic threats, and diseases. The response scale ranged from 1 (strongly oppose) to 5 (strongly support); higher means indicate greater support for monitoring. An aggregate measure of support was created by averaging the ten potentially monitorable activities.

A univariate ANOVA with mean strength of support as the dependent variable and the demographic variables as independent variables was conducted. Average level of support differed significantly by gender (*F*(2, 3,066)=19.81, *p*<0.001; males: *M*=3.65 *SD*=0.75; females: *M*=3.76 *SD*=0.75) and age cohort (*F*(2, 3,066)=6.40, *p*=0.002; youngest: *M*=3.69 *SD*=0.79; middle: *M*=3.70 *SD*=0.77; oldest: males: *M*=3.77 *SD*=0.71), but not income, residency, or education. The pairwise differences between age cohorts were non-significant (18-44 years, *M*=3.69; 45-64 years, *M*=3.71; 65-85+ years, *M*=3.76).

#### Support for Monitoring Specific Locations

Respondents indicated which locations they would want to be monitored out of seven options (e.g., the parts of the city, certain neighborhoods, prisons). One option was to <u>not want</u> any location monitored; another option was to <u>want the entire city</u> monitored. For those who choose neither of these two options, the most commonly chosen location for monitoring was schools (26%) and the least common location was houses (15%). Comparisons by demographics were made for the two options (no locations, all locations), as nearly 90% of the sample selected one or the other. Overall, 76% of the sample endorsed the desire for the entire city to be monitored. There were no differences between men and women, whites and minorities, age cohorts, income, or education levels. However, those living in urban areas endorsed monitoring the entire city at a higher percentage than those living in suburban areas (79% vs. 74%, respectively; Fishers Exact test=0.006).

On the other hand, more people living in the suburbs (n=250, 11.6%) would not support monitoring of any of the locations compared to people living in urban areas (n=76, 8.2%; Fisher’s exact test=0.005). Age cohort was associated with the percent of people not supporting monitoring of any of the locations (Chi-square (df=2)=7.35, p=0.025). The middle-aged cohort (n=131, 12.6%) was less supportive than the young age cohort (n=69, 10.2%) and the older age cohort (n=126, 9.2%) of monitoring any locations. Respondents (men: n=96, 10.2% and women: 226, 10.7%; Fisher’s exact test=0.37) did not differ on not supporting monitoring of any of the locations. White and minority respondents (white: n=271, 10.5% and minorities: 55, 11%; Fisher’s exact test=0.38) did not differ on not supporting monitoring of any of the locations. There was no association of amount of education and the percentage of people not supporting any of the locations (Chi-square (df=2)=0.08, p=0.96); level of no support ranged from high school (n=66, 10.3%), graduate school (n=47, 10.4%) to college (n=213, 10.7%). There was no association of income and the percentage of people not supporting any of the locations (Chi-square (df=2)=0.92, p=0.63); level of no support ranged from lowest (n=122, 10%), highest (n=82, 10.6%) to middle (n=122, 11.2%).

#### Prohibiting Monitoring of Certain Locations

Respondents indicated which, if any, of the seven types of locations they would prohibit monitoring (e.g., houses of worship, elderly care facilities, truck and rest stops), with an option to support monitoring (i.e., prohibit none) of all categories. Most respondents (67%) endorsed no prohibition to <u>locations being monitored</u>. For those who did not chose this option, the most common category of location respondents wanted to be prohibited from monitoring was personal residencies (27%) and the least common category to be prohibited was educational settings (15%). Comparisons by demographics were made for the option, prohibit none. There was no association of race to choosing to prohibit none (Fisher’s Exact test=0.11)

**Gender** was associated with choosing to prohibit none. Male respondents (36%) were more likely than female respondents (31%) to choose to prohibit none (Fischer Exact test=0.007). **Age cohort** was associated with choosing to prohibit none (Chi square (df=2)=7.25, *p*=0.03); the youngest cohort was less likely (25%) to prohibit none compared to the middle cohort (33%) and the older cohort (31%). **Education** level was associated with choosing to prohibit none (Chi square (df=2)=13.68, *p*=0.001); those with high school education (28%) were less likely to prohibit none than those with at least some college (33%) or graduate school (38%). **Income bracket** was associated with choosing to prohibit none (Chi square (df=2)=14.23, *p*<0.001); those in the lowest income bracket were less likely to prohibit none (30%), compared to the middle-income bracket (32%) and highest income bracket (38%). **Residency** was associated with choosing to prohibit none. Suburban dwelling respondents (34%) were more likely than urban dwelling respondents (29%) to choose to prohibit none (Fischer Exact test=0.007). However, respondents living in urban areas endorsed monitoring the entire city at a higher percentage than those living in suburban areas (79% vs. 74%, respectively; Fishers Exact test=0.006).

#### Privacy Attitude Questionnaire

To explore whether demographics predicted variance in the PAQ subscales (exposure, monitoring, personal information, and protection), four stepwise linear regression models were built with demographic variables as predictors. A lower score indicates a higher level of privacy concern within each factor. Note, rather than binning the demographic variables, the full range of options for each variable was used. The PAQ can thus assess respondents’ privacy boundaries within public services, such as a municipal sewer system, or of wider community monitoring.

A significant regression equation was found (*F(*3, 3078)=9.26, *p*<0.001), with an *R*^*2*^ =0.04 for the PAQ factor of exposure. Education level, income, race, and residency were significant predictors. Respondents’ predicted *exposure* score was equal to 26.85 – Age Range (0.60) + Education (0.30) – Race (0.24) + Household Income (0.08). Age Range was measured as 1=under 18, through 9=85 or older (each range was 10 years, except 2 which was 18-24 years). Education was measured as 1=less than high school, 2=high school graduate, 3=some college, 4=2-year degree, 5=4-year degree, 6=professional degree, 7=doctorate. Race was measured as 1=White, 2=Black/African American, 3=American Indian / Alaskan Native, 4=Asian, 5=Native Hawaiian/ Pacific Islander, 6=other, 7=Multiple Races. Household income was measured as 1=less than $10,000 through 12=more than $150,000, with each income range equal to $10,000.

A significant regression equation was found (*F*(2, 3080)=13.38, *p*<0.001), with an *R*^*2*^ =0.009 for the PAQ factor of protection. Education level and race were significant predictors of protection. Respondents’ predicted *protection* score was equal to 30.99 –Education (0.16) + Race (0.28).

A significant regression equation was found (*F*(4, 3078)=15.66, *p*<0.001), with an *R*^*2*^ =0.02 for the PAQ factor of personal information. Education level and race were significant predictors of personal information. Respondents predicted *personal information* score was equal to 24.38 + Education (0.18) - Race (0.32) + Household Income (0.09) – Gender (0.92). Gender was measured as 1=Male, 2=Female, 3=Other. The option “I’d prefer to no answer” was coded as missing and excluded.

A significant regression equation was found (*F*(4, 3078)=15.40, *p*<0.001), with an *R*^*2*^ =0.02 for the PAQ factor of monitored. Education level, gender, age range, and household income were significant predictors of monitored. Respondents predicted *monitored* score was equal to 29.38 - Education (0.58) + Gender (0.75) + Age Range (0.12) – Household Income (0.05).

#### Predicting Support for Monitoring of 10 Activities

A linear regression equation was constructed using a stepwise approach. The predictor variables were entered into the model as follows: mean score of awareness of public health activities, the four PAQ mean subscale scores (exposure, monitored, privacy, and personal information), and the six demographic variables: gender (1=males, 2=females), age group (1=youngest, 2=middle, 3=oldest), race (0=minority, 1=white), income (1=lowest, 2= middle, 3=highest), residency (1=urban, 2=suburban), and education (1=high school, 2=college, 3=graduate school).

A significant regression equation was found (*F*(11, 3055)=32.09, *p*<0.001), with an *R*^*2*^ =0.32 for strength of support for monitoring the 10 activities. Awareness, monitored, protection, personal information, gender, and age cohort remained as significant predictors (*ps*<0.05); exposure, race, education, income bracket, and residency were excluded (*ps*>0.05).

Respondents predicted *Strength of Support* score was equal to 1.37 - Awareness (0.09) + Monitored (0.35) + Protection (0.13) + Personal Information (0.11) + Gender (0.11) + Age Cohort (0.04).

## 4. Discussion

In this study we used a national public opinion survey to understand the public’s perceptions regarding what, where, and privacy concerns in supporting various public health wastewater surveillance activities. We found the prevalence of awareness of wastewater monitoring across the United States was low, but even lower than awareness of restaurant inspections, hotel and motel inspections, and drinking water and public pool water quality testing. Respondents more strongly supported sewer monitoring for terroristic threats, toxins, and disease and indicated the least support for lifestyle behaviors, healthy eating, and mental illness monitoring. In regard to the scale of surveillance, more respondents supported surveillance at a city level over households or business level scales. Our results are consistent with the guidelines by Hall et al. [9] and Scassa, Robinson, and Mosoff [13] which suggest that community wastewater monitoring is generally acceptable, but when monitoring is conducted at smaller scales such as workers, prisoners, and students, it may elicit more concerns. Our national survey results also parallel an earlier study which was focused on views within only Kentucky which showed more public support for wastewater measurements in the largest areas (>50,000 households) [20].

Croft et al. [3] studied both illicit and prescribed neuropsychiatric drugs in wastewater, uniquely spanning choice activities and mental health. Assessing mental health through sewer monitoring, using stress hormones as a quantitative measure, offers an opportunity to highlight the needs and bring more advocacy to fence-line, low-income, or other communities that struggle with environmental justice. However, these privacy concerns of individuals versus a community should be balanced with the real and valid concerns that sewer monitoring could be used as a tool for surveilling and administering punishment or stigmatization upon a community. For example, identifying evidence of illicit drug use within sewers could resulting in negative outcomes for neighborhoods. Alternatively, when sewer monitoring results are made publicly available, it allows individuals and groups, such as those with pre-existing conditions or those who are immunocompromised, to have additional knowledge to assess risk before deciding about participation in public activities. Pertinently, because WBE is currently unregulated, and as the complexities of North Dakota’s proposed legislation of House Bill 1348 [16] shows, who the public could approach about privacy concerns for wastewater surveillance in their city or county remains ill-defined.

The limitations of sewer monitoring’s application in regard to privacy for public health should be acknowledged; WBE is best established when utilizing existing piped wastewater infrastructure. This type of infrastructure covers approximately 85% of the United States population [21] in mostly urban areas, thus allowing a degree of anonymity with a homogenous wastewater sample from many individuals. This is where our survey results show the largest public support. Yet, the remaining 15% of the United States population [21], dominantly rural areas containing more septic tanks or straight pipes or outlier high-income households with large land holdings away from urban centers, would have less individual household privacy in WBE approaches and our survey respondents more often thought this should be prohibited from monitoring.

Although our national survey found high levels of respondent agreement in acceptance for at least some community areas being monitored for wastewater, our work can also guide targeted education programming where public acceptance or concern is comparatively lower. In wastewater reuse, negative public opinion has been found to be driven by pathogen disgust [22]. Yet, wastewater contains more than harmless discarded genetics, and increased use of wastewater monitoring appears to be a part of the future of public health and pandemic preparedness. As the field of WBE continues to build capacity, and with no clear governance on this work, sewer utility providers, public health, environmental health, and the public need to ensure unified support while balancing the need to prevent unethical wastewater monitoring. The results of our study show that even though awareness of wastewater monitoring was low, the guard rails of what was and was not acceptable to monitor were clear and could guide initial policy regulation.

## 5. Limitations

This study did not include a random sample. Our respondents tended to be older women and may represent a participant self-selection bias toward interest in public health surveys and access to internet, in itself an indicator of wealth and access to information. Finally, the results are focused on the United States and further research is needed to gather public perceptions regarding acceptance of wastewater used for community health monitoring globally.

## 6. Conclusion

Using an online survey distributed to a representative sample of adults in the Unites States, we investigated the public’s perceptions regarding what is monitored, where monitoring occurs, and privacy concerns related to wastewater monitoring as a public health surveillance tool. The results suggest that the majority of respondents supported WBE when it is used for public health monitoring, but within some bounds. Being younger in age and urban dwelling were associated with support of wastewater monitoring, compared to older, suburban dwellers. The most important finding of this work may be the absence of a large nationwide concern regarding wastewater being a privacy violation when forming future policy regulation of wastewater monitoring as a public health surveillance tool; and in areas where public acceptance or concern is comparatively lower our results suggest guided targeted education programming.

## Data Availability

All data produced in the present study are available upon reasonable request to the authors

## Authors’ contributions

Conceptualization: ASL and TS; Methodology: TS; Formal analysis: ASL; Writing-original draft preparation: RHH; Writing-review and editing: ASL, RHH, LBA, HDN, TS; Supervision: TS; Project administration: TS. All authors have read and agreed to the published version of the manuscript.

